# Use of a digital application to enhance communication and triage between care homes and National Health Service community services in the United Kingdom: a qualitative evaluation

**DOI:** 10.1101/2023.03.02.23286669

**Authors:** Siân Russell, Rachel Stocker, Zoë Cockshott, Suzanne Mason, Jo Knight, Barbara Hanratty, Nancy Preston

## Abstract

Recent years have seen a rise in digital interventions to improve coordination between care homes and NHS services, supporting remote sharing of data on the health of care home residents. Such interventions were key components in the response to the COVID-19 pandemic. This paper presents findings from the qualitative component of an evaluation of an implementation of the HealthCall Digital Care Homes application, across sites in northern England. The implementation commenced prior to the pandemic and continued throughout.

Semi-structured, qualitative interviews were held with stakeholders. Interviews were conducted remotely (October 2020 -June 2021). Data were analysed via a reflexive thematic analysis then mapped against Normalization Process Theory (NPT) constructs (coherence, collective action, cognitive participation, and reflexive monitoring) providing a framework to assess implementation success.

Thirty-five participants were recruited: 16 care home staff, six NHS community nurses, five relatives of care home residents, four HealthCall team members, three care home residents, and one local authority commissioner. Despite facing challenges such as apprehension towards digital technology among care home staff, the application was viewed positively across stakeholder groups. The HealthCall team maintained formal and informal feedback loop with stakeholders. This resulted in revisions to the intervention and implementation. Appropriate training and problem solving from the HealthCall team and buy-in from care home and NHS staff were key to achieving success across NPT constructs.

While this implementation appears broadly successful, establishing rapport and maintaining ongoing support requires significant time, financial backing, and the right individuals in place across stakeholder groups to drive implementation and intervention evolution. The digital literacy of care home staff requires encouragement to enhance their readiness for digital interventions. The COVID-19 pandemic has pushed this agenda forward. Problems with stability across the workforce within care homes need to be addressed to avoid skill loss and support embeddedness of digital interventions.

**What is known about this topic?:**

- Improving healthcare delivery in UK care homes is a health policy priority.
- Digital interventions designed to enhance the referral process between care homes and NHS services and improve the healthcare delivery in care homes have become increasingly common in the UK. The HealthCall Digital Care Homes application is one such intervention.
- These interventions and their implementations require evaluation to ensure that they operate as intended, function coherently and are considered appropriate and legitimate to the care home setting.

**What this paper adds?:**

- The HealthCall Digital Care Homes app is a feasible, appropriate and legitimate intervention for referral, triage and health care support for non-urgent health care needs of care home residents.
- The ongoing involvement of end users in further developing the intervention, and the level of monitoring and support provided by the implementation team appears to be key to the implementation’s success.
- The digital preparedness of UK care homes is limited. Ensuring that care homes are digitally enabled, with a digitally literate workforce, should be a policy and research priority.

## Background

Residents in long-term residential and nursing care homes have complex health and social care needs. This population has a high degree of multimorbidity, disability and frailty, with impaired cognitive and behavioural functioning (Gordon et al., 2014), which has increased over the past 20 years (Barker et al., 2021). Estimates indicate that emergency admissions and accident and emergency attendances among care home residents are 40-50% higher than the general population ≥75 years (Smith, 2015). Of such admissions ∼50% could be avoided (Harrison et al., 2016). Such complexity can place strain on care homes and the community NHS services that support them.

Improving the quality of healthcare provision in care homes is a priority for the NHS and adult social care (BGS, 2011, 2021). Digital technologies to support communication between care homes and NHS services could support this goal (BGS, 2021). Such interventions use smart devices for the transfer of data such as vital signs observations for the calculation of Early Warning Scores. The National Early Warning Score (NEWS) has been a common component of digital interventions within care homes and community NHS settings (Brangan et al., 2018; Hodgson et al., 2022; Russell et al., 2020; Scott et al., 2019; Stocker et al., 2021) and is being implemented in other countries including Norway (Steinskog et al., 2021). Such interventions have met with some success, including improving communication between health care services and care homes, and instilling confidence in care home staff (Hodgson et al., 2022; Oung, 2021; Scott et al., 2019; Stocker et al., 2021) though the complexity of the care home setting may present barriers (Russell et al., 2020). The need for remote communication between care homes and services produced by the COVID-19 pandemic has led to increased use of and support for digital interventions within care homes (BGS, 2021; Chu et al., 2021; Edelman et al., 2020; Stocker et al., 2021).

### HealthCall Digital Care Homes Application

HealthCall is a collaboration of seven NHS Foundation Trusts across the Northeast of England and North Cumbria (NHSHealthCall, [online]). It focuses on producing digital solutions to health care challenges. One such solution is the Digital Care Homes application, designed to enhance reporting of non-urgent referrals. The app aims to shorten referral times between care homes and NHS services, with the transfer of relevant information through the app as opposed to care home staff waiting in a queue on the phone. Through the app care home staff can record:

1. vital signs to calculate NEWS
2. contextual information (free text format) including “soft” signs of deterioration (changes in behaviour, mood, sleep, appetite, toileting).

The reporting structure uses the Situation, Background, Assessment, Recommendation (SBAR) tool (Leonard et al., 2004), designed to promote organised communication of necessary, contextual information about patients. The referral is reviewed by a clinician at a Single Point of Access (SPA), who requests further information from the care home or triages to an appropriate service. The care home is notified of action taken. Senior carers are most likely to use the app as part of their role.

Care homes are given a digital device and in-house training from HealthCall’s local Clinical Trainers. Training covers the app’s purpose, using the digital device, taking vital signs and what to record on the SBAR tool. App use is monitored by HealthCall. Members of community NHS teams, such as community nurses, also communicate with HealthCall and can support care homes with the app.

This paper concerns the qualitative component of an evaluation of the Digital Care Homes application’s implementation into residential and nursing care homes in Northern England.

### Patient and Public Involvement (PPI)

Patients and the public were involved at the idea generation stage, confirming that research to improve care for care home residents is viewed as a priority. A PPI panel was established. Research questions, topic guide, study design, findings and dissemination strategies, were discussed via a series of online meetings.

## Methods

A phenomenological approach was undertaken. Methods of qualitative inquiry were used, seeking to gain in-depth data about participants’ experiences of and views towards the intervention and its implementation.

### Identification and sampling

Relevant stakeholders included the local HealthCall team, local authority staff involved in the implementation, care home staff and residents, relatives of residents, and community NHS staff.

Care home and NHS staff were recruited using purposive sampling aiming for variety in terms of care home size and type, and type of clinician. Convenience and snowball sampling were then used to increase sample size. Residents and relatives were recruited using convenience and snowball sampling.

### Recruitment

A member of the local HealthCall team made initial contact, via email, with their colleagues, a Local Authority Commissioner who worked on the implementation, care homes and NHS services on behalf of the research team. They provided a brief description of the evaluation and contact details for the research team who then followed-up on this initial contact.

Care home staff introduced the evaluation to residents who had capacity to provide informed consent. Relatives were sought using a short advert through online community networks. Interested stakeholders were given a PIS and offered the opportunity to ask questions about the evaluation and their participation. PIS were adapted for each stakeholder category. If the wish to participate was upheld a suitable time for data collection was arranged. As the evaluation was conducted remotely, informed consent was secured electronically or verbally, meeting Health Research Authority principles for remote consent.

### Data Collection

Data were collected between November 2020 and July 2021 via semi-structured interviews, one-on-one, dyadic, and small group. Interviews were conducted online using video conferencing platforms or by telephone. The topic guide was developed based on similar evaluations conducted by members of the research team (Russell et al., 2020; Stocker et al., 2021) and evaluation aims. As is typical for semi-structured interviews, the topic guide acted as an aide-mémoire rather than being rigidly followed. Interviews were audio-recorded with permission from the participant(s). Data collection ceased once data sufficiency (Dey, 1999) was realised.

### Data Analysis

An initial phase of analysis followed the six-phase process for reflexive thematic analysis (TA) (Braun & Clarke, 2019, 2021): familiarisation, generating initial codes, searching for themes, reviewing themes, defining themes and analysis write-up.

Tentative initial codes were discussed among the team based on relevant previous work. Rather than “bracketing” these potential codes they were included in the initial phase of coding and removed if not relevant, thereby informing, but not leading, early analysis.

In reflexive TA coding is “fluid, organic, and recursive” where codes can “expand, contract, be renamed, split apart … collapsed together … and even be abandoned” (Braun & Clarke, 2021). This reflects our approach to this process. RS, ZC, and SR led the analysis, independently coding transcripts, and meeting regularly to review codes and collaborate on the production, reviewing and defining of themes. Developing analysis was discussed with the wider qualitative team, NP and BH, at various stages of data collection and analysis, supporting the sense making work of analysis (Braun & Clarke, 2019, 2021).

Data were then considered against Normalization Process Theory (NPT) (May & Finch, 2009; Ross et al., 2019) constructs of coherence, collective action, cognitive participation, and reflexive monitoring to provide a framework to evaluate implementation success (see Table 3). Similar approaches have been undertaken elsewhere (Asiedu et al., 2019; Ross et al., 2019; Russell et al., 2020). Conducting a TA prior to considering NPT constructs ensured that the voices of participants were accounted for, and analysis was not driven solely by an existing framework.

## Results

### Participants

Thirty-five participants were recruited (see Table 1). The majority, sixteen, were care home staff. Six of the eight care homes were residential only, two were independently run and they varied in size. Six NHS community nurses, three residents and five relatives participated. The relatives were not related to residents interviewed. Five participants directly involved in implementation were interviewed: four from the local HealthCall team and one Local Authority Commissioner.

**Table 1:**
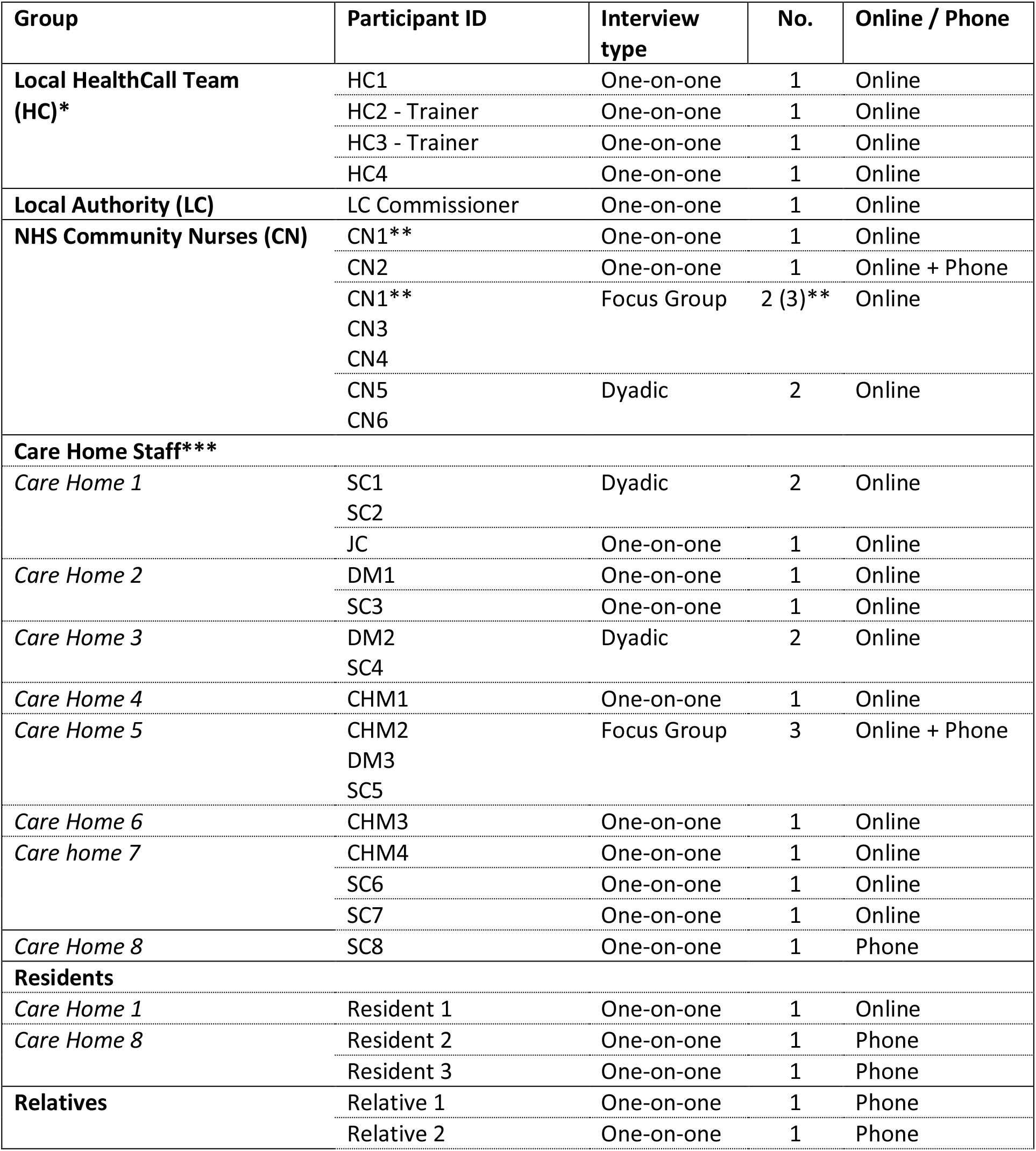
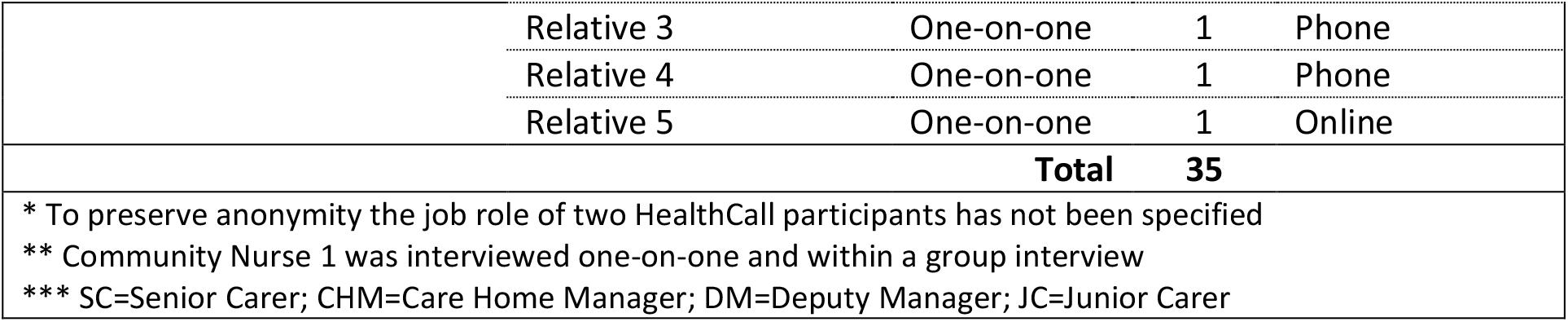
Participants.

| Group | Participant ID | Interview type | No. | Online / Phone |
| --- | --- | --- | --- | --- |
| **Local HealthCall Team (HC)\*** | HC1 | One-on-one | 1 | Online |
|  | HC2 - Trainer | One-on-one | 1 | Online |
|  | HC3 - Trainer | One-on-one | 1 | Online |
|  | HC4 | One-on-one | 1 | Online |
| **Local Authority (LC)** | LC Commissioner | One-on-one | 1 | Online |
| **NHS Community Nurses (CN)** | CN1** | One-on-one | 1 | Online |
|  | CN2 | One-on-one | 1 | Online + Phone |
|  | CN1** | Focus Group | 2 (3)** | Online |
|  | CN3 |  |  |  |
|  | CN4 |  |  |  |
|  | CN5 | Dyadic | 2 | Online |
|  | CN6 |  |  |  |
| **Care Home Staff\*\*\*** |  |  |  |  |
| *Care Home 1* | SC1 | Dyadic | 2 | Online |
|  | SC2 |  |  |  |
|  | JC | One-on-one | 1 | Online |
| *Care Home 2* | DM1 | One-on-one | 1 | Online |
|  | SC3 | One-on-one | 1 | Online |
| *Care Home 3* | DM2 | Dyadic | 2 | Online |
|  | SC4 |  |  |  |
| *Care Home 4* | CHM1 | One-on-one | 1 | Online |
| *Care Home 5* | CHM2 | Focus Group | 3 | Online + Phone |
|  | DM3 |  |  |  |
|  | SC5 |  |  |  |
| *Care Home 6* | CHM3 | One-on-one | 1 | Online |
| *Care home 7* | CHM4 | One-on-one | 1 | Online |
|  | SC6 | One-on-one | 1 | Online |
|  | SC7 | One-on-one | 1 | Online |
| *Care Home 8* | SC8 | One-on-one | 1 | Phone |
| **Residents** |  |  |  |  |
| *Care Home 1* | Resident 1 | One-on-one | 1 | Online |
| *Care Home 8* | Resident 2 | One-on-one | 1 | Phone |
|  | Resident 3 | One-on-one | 1 | Phone |
| **Relatives** | Relative 1 | One-on-one | 1 | Phone |
|  | Relative 2 | One-on-one | 1 | Phone |

|  |  |  |  |
| --- | --- | --- | --- |
| Relative 3 | One-on-one | 1 | Phone |
| Relative 4 | One-on-one | 1 | Phone |
| Relative 5 | One-on-one | 1 | Online |
| **Total** |  | **35** |  |
| * To preserve anonymity the job role of two HealthCall participants has not been specified |  |  |  |
| ** Community Nurse 1 was interviewed one-on-one and within a group interview |  |  |  |
| *** SC=Senior Carer; CHM=Care Home Manager; DM=Deputy Manager; JC=Junior Carer |  |  |  |

Twenty-four interviews were one-to-one. Community Nurse 1 was interviewed individually and participated in a group interview. Twenty-two participated via video conferencing.

## Findings

The TA process resulted in three themes Theme 1: “It’s a bit like anything new”: Anticipated, unexpected and implicit challenges of implementation, Theme 2. Communication and Training and Theme 3. Efficiency and Appropriacy, as detailed below. Exemplar quotes are presented in Table 2.

**Table 2:** Themes and sub-themes and exemplar quotes.

| **Theme 1 "It's a bit like anything new": Anticipated, unexpected and implicit challenges of implementation** |  |
| --- | --- |
| *Digital skills and confidence* | *...all of [...] made an assumption that all people in care homes are going to be happy to use a digital technology. Even though it's no different to some of the phones they're using, some are very, very afraid of digital technology*<br>**HC3 - Trainer**<br>*... the main issue we've found is the lack of IT skills in care homes [...] even now I think we probably are still quite astounded by how poor they are*<br>**HC1**<br>*[SC4's] a technical wizard ... She keeps me right. I'm the technophobe [...] I let [HealthCall 3 - Trainer] show [SC4] and then [SC4] showed me because I'm very slow with technology [...] But no, I'm getting there.*<br>**DM2** |
| *Practices, Habits and Cultures* | *... rather than doing the easy thing and picking up the telephone [...] changing that habit [...] There's a lot of information you have to input that seems a bit much when you're just trying to get someone out to have a look at someone [...] what their blood pressure is and all the rest of it [...] we need to do this, this and this, before we actually go on the iPad to put it into the HealthCall system.*<br>**CHM1**<br>*...it's a bit like anything new I think. It takes a little while to get it embedded and entrenched and become custom and practice and used to.*<br>**HC2 – Trainer** |
|  | *...not an extra job, it's an 'instead of' job* **HC2 – Trainer**<br>*...they just want to speak to a human [...] That's a human response [...] We thought it was really well embedded but actually when there's a pandemic it's a phone call...* **CN1 (one-on-one interview)**<br>**CN6:** *... they've worked out what makes us tick [...] They manipulate. [...] we've made them quite needy because we're there as soon as they need us. [...] sometimes they don't do the observations and one of the excuses is they say the patient won't allow them and then obviously when we go there they do allow you to do observations [...]*<br>**CM5:** *Yeah they'll make an excuse as to why they haven't done [the observations] but then tell you all sorts of other things that will make you turn up [...] if you had the observations you could actually say, 'Well this person's temperature isn't up, the blood pressure isn't that low, [...] the blood pressure is normal,' but if they don't do that then you're going on what they say the patient is like, which could be at times fabricated.*<br>**Interviewer:** *Do [residents] mind having their vital obs. taken? Sounds like it was something you were doing...*<br>**SC7:** *Yeah it was something we would do anyway so not really. Obviously it can be difficult if a resident has dementia and they don't want them doing. That's fine so there is an option to click no observations but then just to put the reason why, so you can put in that the resident refused...* |
| Technology and Operations | *[...] we do have problems with one of our tablets [...] in that we can't access the camera [...] you can take wound assessments and take pictures and we can't do that. We have had support though. There's a gentleman who comes and sees us regularly and he tried to fix it I don't know how many times but it hasn't worked. I would say that's probably the only technical issue we've had.* **CHM1**<br>**Interviewer:** *[...] What is better for them in picking up a phone?*<br>**HC3 - Trainer:** *Usually it is access to the internet or access to the technology. So a lot of homes have very poor internet [...] So even though they've got the smart pads, then if they were in a certain area of the home then they can't access the internet anyway. Quite a lot of homes operate so that the care staff do not use the office space, so they don't have access to the desk top which could be another alternative to using our app [...] it does tend to be the convenience of using it. [...] I'll give you an example, one of our homes had a 33% use against their population, and once we gave them a phone that they could drop into their pocket and it was 4G so it was connected to the 4G rather than their home internet, last month's figures was 130% usage – so they used it for everything. [...] So that's what we've got to work on – finding the quickest route and the easiest route for each individual care home because no one size fits all.*<br>*[...] from a strategic point of view we have bombarded the homes with equipment and actually we were just adding a load of burden [...] We've given them tablets, NHS England or X [...] are offering them iPads left, right and centre. [...] actually what do the care homes need? We're all assuming that they need this stuff [...] but actually we're potentially creating a massive burden for their IT people and their own infrastructure [...] we don't want it to be at the point where we've got a tablet for HealthCall, a tablet for this, a tablet for this... [...] a lot of this is being done knee-jerk reaction to covid obviously- [...] **HC1*** |
| **Theme 2: Communication and Training** |  |
| *Relationships and support* | *They would rather just pick up a phone up [...]we're trying to tackle that [...] we'll get a message back from the Single Point of Access to say they've received the call, they're taking the care workers name now and then that way we can pass that on to the care home managers and say does this person need some more training? [...] we'd be sending hints and tips through via email – group emails with the care homes just reminding them all when baseline obs. were needed and when they weren't [...] HC4*<br>*[...] either myself, [or other team members] would kind of make the initial approach to the home and basically explain as to what we were doing [...] where it all came from as a joint exercise between the CCG, the Trust, ourselves [...]then we basically arranged a time to go and do some training focussed mainly on either the manager or the seniors or both [...] then schedule in a revisit the following week just by way of support [...] give them our mobile numbers, our email addresses [...] and give them a ring [...] then follow that up with regular visits. Once they're established on it once a month per home [...] if homes are struggling post roll out they'll need a lot more support... HC2 - Trainer*<br>*Any concerns that we've got they're very quick at responding and fixing the problem. SC6*<br>*...we ask them why they haven't used the [app]. Once we know why they haven't and they do tell us, we let the [local HealthCall] team know if there's any problems with training or passwords and they pick that up directly with the care homes... CN2*<br>**CN4:** *... Some of them use [HealthCall] better than others, some forget [...] you kind of go back in and say, 'Can you start using it again?' and they do.*<br>**Interviewer:** *So, in that case would you say that you encouraged the care homes to use it as much as possible?*<br>**CN4:** *Yeah definitely.*<br>**Interviewer:** *Was that something [...] you arranged with HealthCall for themselves or was that because you think it's such a good intervention?*<br>**CN1:** *[...] we knew that it was an intervention that needed exploring [...] we went to a couple of meetings [with the implementers] we had the opportunity to put that human side to it instead of somebody from above telling us what we needed to be asking we were saying, 'Well we wouldn't ask in that way,' [...] so we had that opportunity to build it which was really good and that was definitely a success.*<br>*What we experienced initially was some resistance from our specialist practitioners who wouldn't accept triages without a full set of observations and being slightly unrealistic about what the care homes could produce. That obviously was discussed [...] we came to an agreement about what could and couldn't and would and wouldn't be accepted. HC3 – Trainer*<br>*[...] sometimes it was just to sit down with the manager and going, 'Are you ok?' and just doing a bit of a welfare check with them [...] We've got good relationships with the managers and [the HealthCall Trainers] were able to just to have that bit of time that a lot of other medical or clinical staff weren't able to [...] HC1*<br>**Interviewer:** *[...] Have you come across [the Digital Care Homes app] at all?*<br>**Relative2:** *No, I haven't. [LATER] I wouldn't have to be told, no. It would be interesting to know I suppose but, so that I could have sat with him for some of them. It would have been good if they did it that way [...] but no I* |
|  | wouldn't mind if they didn't say, whatever they have to do to help people is fine. **Relative 2** |
| Appropriacy of training | [...] when it was first launched we were involved in the training of the staff members [...] so [care homes] had sort of a friendly face... **CN1**<br>**DM2:** [HC2-Trainer] told us how it was set up on the actual pad. Then showed us what to do with our usernames, our passwords and then gradually we got down to well this is how you get in touch with the community matrons. This is how you get in touch with the practice managers [...] this kind of thing [...] Initially [HC2-Trainer] came quite a bit but it was like maybe trying to catch people on different shifts as well and ensuring that we all were singing from the same song sheet [...] [HC2-Trainer] was very patient. No question was silly [...]<br>[...] In the COVID roll out, the speedy roll out, we mainly cut it down to the digital technology only. If we had nurses in the nursing homes we asked them to train the clinical skills [...] Quite a few of our homes already had carers that had been through their company taught clinical skills [...] But the difficulty with COVID was the getting the access to the staff so we did a lot of remote training [...] training in a car park [...] But because of that we did miss some of the more resistant staff because it gave them a chance to hide [...] We are now mopping those people up now [...] **HC3 - Trainer** |
| Application and implementation evolution | **SC6:** ... they came to the home and did sessions with all our seniors and managers and stuff and then me and [colleague 1] went to a meeting with the head of the [community] nurses, the ones who were – what are they called...?<br>**Interviewer:** At the Single Point of Access? [...]<br>**SC6:** Yeah. It was like people from like different care homes were there and it was really interesting. It was just a bit of a catch up of how everyone's finding it. Was there any concerns? [...]<br>[...] we've got a generic email folder where the care homes can send any queries in if the staff's got issues with log-ins. I can help them out with that. If they've got new residents or if they've got leavers all of that needs to be constantly updated otherwise the system won't work and that's getting busier and busier [...] to cope with. **HC4** |
| **Theme 3. Efficiency and Appropriacy** |  |
| Enhancing resident care and improving efficiency | We click the person's name and then it will ask you what are you concerned about? Have you got observations? And it just says blood pressure, temp, oxygen levels, things like that and you just fill in the boxes, the questions. Have there been changes? What have you done to help already? Things like that so and then you submit and it's that easy. **SC7**<br>**Interviewer:** [...] would you want to go back to life without it?<br>**SC6:** No [...] I mean you could be in a queue and you could be number nine in that queue for an hour and a half and you've got other things to be doing [...] That patient could be getting progressively worse whereas if you've got the app you can just put the observations straight on it and it'll go directly [...] It just cuts out the time.<br>[...] it was a reduction of two admissions per month from the care homes that were in [the pilot] [...] 'Right so times that by 12 months by [all] care homes,' and you're talking mega savings [...] the feedback from [...] the frontline staff who were using the system [...] how much time it saved them [...] They were able to spend more time with the residents... **LAC** |
|  | *[...] because of COVID the [local HealthCall] team pushed through access for all of the care homes quicker, they finished much faster than they were originally planned to do it then uptake was really, really good. **CN2***<br>*Oh yes. I got a wonderful examination [...] on the computer, yes [...] I got peace of mind in a very short time. **Resident1*** |
| Accountability and legitimacy | **HC2 - Trainer:** That's where the kind of safety net is. They've [SPA] got a 15 minute response time.<br>**Interviewer:** Okay so that's a 15 minute response time from when they get that referral through [...]?<br>**HC2 - Trainer:** Yes [...] you get obviously a full case history and everything with [digital health record]. They can get the NEWS score, they'll get all the relevant information that comes through from the app and they can sort of align that to [digital health record]. [...] if you're not happy with the obs. [...] just phone them you know [...] Our understanding is once that referral comes through we've then got clinical responsibility for that patient in terms of the [SPA] side of things ...<br>*[...] "they're just not right" and being just not right is a clinical sign for elderly [...]*<br>*[...] as a registered nurse I'm, I don't think it's [NEWS] something we should be using as a diagnostic tool [...] the actual scoring on it is too prescriptive for the co-morbidities within care and nursing homes [...] NEWS itself, is a very useful triage tool [...] and at HealthCall we don't use NEWS for a diagnostic we only use it as a guide to how ill the person could be and take the other things into consideration. **HC3 - Trainer*** |
| Upskilling | *I think the app itself has developed people's technological skills as well. Everyone in here was a little bit scared about using it to start off with but it's really straight forward but I think yeah staff are learning [...] when they read someone's blood pressure they're going, 'Oh no that's a bit high,' or 'That's a bit low,' so they are starting to understand that. **CHM4***<br>*[...] descriptive language is vastly improved. They are looking at the wider picture [...] They put things like, 'Has just finished antibiotics. Still got a chest infection' [...] we're getting a lot more detail in there now... **CN1, One-on-one Interview*** |

**Table 3:** Findings in relation to NPT Constructs.

| Table 3: Findings in relation to NPT Constructs |  |
| --- | --- |
| **Core Construct: Coherence**<br>*Process of sense-making and understanding that individuals and organisations go through in order to promote or inhibit the routine embedding of a practice to its users. These processes are energized by investments of meaning made by participants.* |  |
| **Differentiation:** Do stakeholders see this as a new way working? | Acknowledged across stakeholders, particularly for care home staff: Referral via app not phone, specific contextual information requested, increased digital and clinical knowledge required.<br>Community nurses discussed how the app altered practice such as approach to prioritising visits. |
| **Individual specification:** *Do individuals understand what tasks the intervention requires of them?* | Stakeholder understanding of the intervention and their role was clear.<br>Problems with digital skills in, and preparedness of, care homes a potential barrier to engaging coherently with the intervention.<br>Disparaging comments about care home staff (two community nurses) suggests the potential for sociocultural barriers to Individual Specification. |
| **Communal specification:** *Do all those involved agree about the purpose of the intervention?* | Stakeholder's all commented on the core aims of the intervention: reducing referral time, improving communication and quality of information, enhancing resident care and reducing avoidable hospital admissions.<br>Care home staff defaulting to the phone to refer and the disparaging comments of two community nurses suggest some undermining of Communal Specification |
| **Internalisation:** *Do all the stakeholders grasp the potential benefits and value of the intervention?* | Achieved across stakeholder groups<br>Stakeholders discussed the upskilling of care home staff, improvements to efficiency and communication, quality of data for triage and prioritising visits, and improvements to resident care<br>The remote link between care homes and community NHS staff enhanced by COVID-19. |
| **Core Construct: Cognitive Participation**<br>*Process that individuals and organisations have to go through in order to enrol individuals to engage with the new practice. These processes are energized by investments of commitment made by participants.* |  |
| **Enrolment:** *Do the stakeholders believe they are the correct people to drive forward the implementation?* | The local HealthCall team appeared confident in their roles within the implementation<br>The Community Nurses participated in activities to support and sustain the implementation, providing training to care home staff and encouraging engagement<br>Care home managers and seniors were supportive of the app |
| **Initiation:** *Are they willing and able to engage others in the implementation?* | The local HealthCall Team and Community nurses were concerned with encouraging and supporting other stakeholders to engage with the intervention, monitoring use and engagement and managing relationships<br>Care home managers and Senior carers were involved in supporting use and engagement within care homes |
| **Activation:** *Can stakeholders identify what tasks and activities are required to sustain the intervention?* | HealthCall identified ways to improve and adapt the implementation over time, ensuring the app and implementation were appropriate.<br>Community nurses supported care home staff and alerted HealthCall to training needs in care homes<br>Habits and issues of convenience could undermine activation. Care homes increasingly utilised the app in daily practice and alerted HealthCall where further support was required |
| **Legitimation:** *Do they believe it is appropriate for them to be involved in the intervention?* | Care home managers and seniors, and community nurses were supportive of the app and their engagement with it appeared to be viewed as a valid part of both care and health care.<br>Onus of responsibility for high level clinical decisions was not placed on care home staff |
|  | Local HealthCall team enthusiastic about their and had the right experience and skills to drive implementation. |
| **Core Construct: Collective Action**<br>*The work that individuals and organisations have to do to enact the new practice. These processes are energized by investments of effort made by participants.* |  |
| **Interactional workability:** *Does the intervention make it easier or harder to complete tasks?* | Improvements to efficiency were often commented upon across stakeholder groups<br>For care home staff the ease and quick referral process was key<br>Community nurses appreciated the steady improvement in information they received. They felt more prepared for visits and could prioritise their visits<br>Poor digital literacy among care home staff had the potential to undermine this |
| **Skill set workability:** *Do those implementing the intervention have the correct skills and training for the job?* | The HealthCall Team had the appropriate skill set and experience for their roles with care home staff commenting positively on their approach to training and approachability<br>Buy-in and support from community nurses and care home managers meant that their skills as implementation allies bolstered the work done by the HealthCall team. |
| **Relational integration:** *Do those involved in the implementation have confidence in the new way of working?* | While the implementation experienced some challenges, views toward the intervention were broadly positive and HealthCall were proactive in dealing with issues that arose<br>The negative views concerning motivations of care home staff in using the app, exposed by two of the Community Nurses, suggest that there may be sociocultural barriers |
| **Contextual integration:** *Do local and national resources and policies support the implementation?* | Current drive to improve health care delivery within care homes, at both local and national level<br>Clinical Commissioning Groups, NHS Trusts and Local Authorities across the UK have sought to implement and evaluate similar digital innovations<br>Government pledge to digitised adult social care by March 2024<br>HealthCall raises a warning that care homes should not be “bombarded” with technology |
| **Core Construct: Reflexive Monitoring**<br>*Informal and formal appraisal of a new practice once it is in use, in order to assess its advantages and disadvantages and which develops users’ comprehension of the effects of a practice. These processes are energized by investments in appraisal made by participants.* |  |
| **Systemisation:** *Will stakeholders be able to judge the effectiveness of the intervention?* | Monitoring of app use by HealthCall<br>Evaluation: Quantitative component examining changes in referral patterns and decisions, rates and nature of hospital admission and discharge. Qualitative component exploring usability and acceptability, impact on referral practices and resident care, indicators of implementation success |
| **Individual appraisal:** *How will individuals judge the effectiveness of the intervention?* | No formal indicators or measures for how individuals appraised the intervention beyond their own personal interactions with the app and other stakeholders<br>This evaluation offered stakeholders the opportunity to express their views |
| **Communal appraisal:** *How will stakeholders collectively judge the effectiveness of the intervention?* | • Steps taken to judge effectiveness were typically “informal” by HealthCall. They maintained a dialog with care homes, community nurses and other stakeholders such as the Local Authority Commissioner, creating a feedback loop<br>• Stakeholders had discussed the intervention within their own teams highlighting successes and problems |
| **Reconfiguration:** *Will stakeholders be able to modify the intervention based on evaluation and experience?* | • The feedback loop created by the relationships between the local HealthCall team and other stakeholders allowed for both the intervention and the implementation to be adapted and to evolve in response to feedback from end users<br>• The involvement of key stakeholders has been ongoing since the development stage<br>• HealthCall also creatively adapted approach to training in response to COVID-19 |

Theme 1. “It’s a bit like anything new”: Anticipated, unexpected and implicit challenges of implementation

The implementation faced challenges anchored on pre-existing skills and confidence of staff, workplace habits and inter-professional dynamics, and problems with the technology.

### Digital skills and confidence

Poor digital skills and limited confidence using digital devices among care home staff was widely acknowledged across stakeholder groups. Older care home staff were viewed as particularly apprehensive. HealthCall participants identified this as a considerable, yet unanticipated challenge. There was an assumption that the general prevalence of smart technology meant staff would be digitally literate. However, the need for digital skills training was viewed as greater than for training on vital signs observations.

Some care home staff mentioned their own lack of digital confidence. This was flagged as a training need that went beyond the Digital Care Homes application.

### Practices, Habits, and Cultures

Care home staff could fall back into pre-intervention practices, particularly using the phone for referrals rather than the app. **CHM1**, while supportive of the intervention and aware that changing behaviour could take time, believed the volume of information required by the app could seem more time consuming than picking up the phone. The local HealthCall team believed such challenges should be anticipated in the introduction of novel interventions. **HC3 - Trainer** stated the need to stress that the app was “*not an extra job, it’s an ‘instead of’ job*” during training to tackle this issue.

Reverting to using the phone to make a referral also occurred as a response to the COVID-19 pandemic. The local HealthCall team assumed this was due to anxiety caused by COVID-19, a view shared by **CN1: “***they just want to speak to a human […] That’s a human response*”. **CN1** felt this questioned how embedded the intervention was prior to the pandemic.

Negative views toward care home staff were expressed by two community nurses. While they acknowledged the challenges within care homes, including staff turnover, and the pressure faced by care home nurses, they characterised some staff as “needy” and “manipulative” in their use of the app. These community nurses also believed that care home staff would make “excuses” for when a resident’s observations could not be obtained. This highlights the potential for problematic relationships between stakeholders. That obtaining observations is not always possible was understood by HealthCall and the option to not put observations into the app was something HealthCall provided.

### Technology and Operations

Issues with the technology could disrupt the use and embedding of the intervention. The technology could occasionally not work as intended, for example, referrals sent but not received. Unreliable Wi-Fi signals in some care homes could restrict app use to certain parts of care homes or to a desktop computer, typically within the manager’s office, rather than a tablet. **HC3 - Trainer** believed app use would decline where the convenience of using it was hampered, an issue that poor Wi-Fi signals created. HealthCall discussed problem solving such issues by installing the app onto smartphones with 4G, expanding online access.

**HC1** warned that care homes have been “bombarded” with digital technology, especially in response to COVID-19, and was concerned that care homes might end up with too many tablets and applications causing a burden to staff and the IT capability of care homes.

Theme 2: Communication and Training

The HealthCall team maintained relationships with NHS services and care homes, working with them and supporting them as required, and adjusting the training and the app due to this collaborative approach.

### Relationships and support

The local HealthCall team discussed the processes involved in developing a rapport with care homes, monitoring use of the app and providing additional support and problem solving, whether this was training or technical support, as required. This work was proactive and involved communication between the HealthCall team, the Single Point of Access, NHS community staff and the care homes, and thus considerable co-ordination.

The local HealthCall team’s relationships with community NHS services could be harnessed to further support the care homes and identify problems. Buy-in from community nurses meant that these stakeholders, who maintain regular contact with the care homes, could encourage engagement with the app. Relatedly, members of the local HealthCall team spoke about having to negotiate expectations and relationships between care homes and NHS services in relation to the intervention. However, the negative views expressed by two community nurses (discussed in Theme 1) suggests these negotiations may be an ongoing requirement for the intervention to function coherently. HealthCall also contacted care homes to check how they were coping with COVID-19, potentially further embedding these relationships.

Relatives of residents were not explicitly familiar with HealthCall. Relatives were ambivalent about whether they should be informed of the intervention. Whether families were informed about or aware of the intervention could differ across the care homes. Relatives felt that families would not mind how healthcare professionals were contacted, as long as residents received appropriate health interventions.

### Appropriacy of training

The HealthCall Trainers had appropriate experience and knowledge for their job such as considerable clinical experience and one had a background in intervention implementation. **HC1 - Trainer** also commented positively on their own HealthCall induction: *I don’t want to blow smoke, but it was the best induction I’ve had*. Some community nurses provided training during the early phases of the implementation which meant the app was introduced to care homes by someone they were familiar with.

Training was delivered to staff within their own care home. Therefore, practice taking vital signs and using the app occurred in a real-world setting. This was followed up with further visits from the Trainers. The approach to training was well received by care homes and care home staff viewed the Trainers as patient and approachable.

HealthCall amended the approach to training in response to the different phases of implementation, the COVID-19 pandemic, and to address poor digital literacy levels amongst care home staff. In response to the pandemic, the implementation was accelerated to ensure all care homes had use of the app for remote monitoring. The Trainers engaged in pragmatic workarounds, delivering training flexibly. This compromised the depth of the training and further resource had to be spent later to “mop up” (**HC2 -Trainer**) staff that had missed the training delivered in the early months of the pandemic.

### Application and implementation evolution

Various participants discussed the app being developed with care homes and NHS staff and acknowledged the feedback loop of ongoing communication between these stakeholders and HealthCall. This meant that the app improved and evolved with insight from end users, enhancing its appropriacy and legitimacy. The language used within the app also evolved and other adjustments, such as allowing care home staff to not report all vital signs if a resident refused, were added.

The perceived successes of the implementation relied on monitoring app use, providing training and technical support, listening to and responding to feedback from various stakeholders. However, **HC4** noted that this created considerable work and was at risk of stretching resources: *all of that needs to be constantly updated otherwise the system won’t work and that’s getting busier and busier […] to cope with*.

Theme 3. Efficiency and Appropriacy

### Enhancing resident care and improving efficiency

Despite issues with digital skills in care homes the view that the intervention was easy to use was common. Care home and NHS staff highlighted the improvements to efficiency afforded by the app, enhancing its legitimacy within this setting. For care home staff, the ease of use and prompt referral process meant they had more time for other tasks and for residents. For community nurses, having vital signs and contextual information about residents in advance of a visit was valuable: *…it helps with your priorities…* **CN1**. The HealthCall team and LA Commissioner also highlighted these benefits.

While the residents interviewed did not appear to be fully aware of the intervention, one resident, **Resident1**, recalled having her vital signs taken and being seen by a medical professional quickly. She described this as giving her “*peace of mind in a very short time*”. **Resident2** mentioned being happy to have her temperature taken every morning. This was mentioned when discussing COVID-19, and as such does not necessarily speak to the app. Relatives were also not fully aware of the intervention. Opinions towards it, as an abstract idea, explained during interviews, were generally positive.

### Accountability and legitimacy

A key element of the app is how accountability has been addressed. While care home staff are responsible for recognising a potential decline in a resident’s health and communicating relevant information within a referral, the SPA acts as a safety net. SPA staff are clinically trained with access to patient records via digital health records, providing them with further information on the resident of concern. If SPA have concerns about the information received from a care home, they can contact the care home for clarification. As the clinical experience of care home staff is variable and often limited, placing the responsibility for interpretation of clinical information and response onto the SPA removes this potential burden of clinical accountability from the care homes.

Care home staff spend considerable time with their residents and typically recognise ‘soft’ signs of deterioration. This pre-existing knowledge was deemed vital for the intervention to perform coherently. This intervention legitimised these skills by placing them on par of importance with more objective observations such as vital signs. The local HealthCall team and community nurses appreciated that NEWS alone is not an appropriate tool for diagnosis, but part of a wider overview of the resident. This recognition prevented the intervention from having an onus on obtaining all vital signs, which is not always possible.

### Upskilling

Stakeholders believed the app improved the digital literacy and capability of care home staff. This may have been influenced by the increased need for remote monitoring and digital communication created by COVID-19. Improvements in communication were noted with participants across stakeholder groups observing that clinical knowledge among care home staff had advanced and the information they provided to support referrals had improved.

### Normalisation Process Theory

Table 3, below, outlines how the findings from the thematic analysis relate to NPT constructs (further detail provided in Appendix S1). Resident and relative stakeholders’ awareness of the intervention was limited and their involvement in the implementation passive as opposed to active. As such the NPT findings relate chiefly to other participants.

## Discussion

The core themes presented above provide a clear narrative around the challenges faced by the ongoing implementation of the Digital Care Homes app, and how those involved have sought to find solutions, harnessing a feedback loop born of the open and positive communication the implementers created. When considered against NPT constructs, the findings confirm that the implementation has been comprehensive and adaptive, with proactive and appropriately skilled individuals driving the implementation, which is appropriate for a complex intervention.

The intervention has broadly been viewed as a legitimate and valuable contribution to care work and health care delivery, achieving buy-in from key stakeholders. This buy-in was informed by positive experiences with the app, chiefly the ease of use and efficient referral time. Community nurses appreciated the additional information they received that enabled them to better prepare for their visits. These positive opinions suggest that this intervention was meeting its aims. Findings from previous work evaluating interventions like the Digital Care Homes app (Russell et al., 2020; Stocker et al., 2021) suggests that, for this kind of intervention to achieve buy-in from care home staff and community NHS services implementation teams should: -

- work *with* key stakeholders in developing the intervention and implementation
- account for the challenging nature of care homes including multiple competing priorities and medically complex residents
- understand that obtaining vital signs from residents is not always achievable and care home staff should not be accountable for clinical decisions.
- respect care home staffs’ ability to recognise soft signs of deterioration and acknowledge the importance of such information
- provide training that is comprehensive with a practical element (taking vital signs on care home residents in-house), delivered by those with appropriate clinical skills and experience with care homes
- monitor engagement and use of the intervention, flagging any further support needs
- provide ongoing support with digital technology and further training.

The development and implementation process undertaken by HealthCall appears to have accounted for these issues.

It should be noted that NEWS was developed for use in acute rather than community settings (RCP, 2017). Concerns have been raised about the use of NEWS with medically complex populations (Grant, 2018; Grant & Crimmons, 2018) questioning its appropriacy for use with care home residents. NEWS is not a standalone tool, and should be considered alongside further contextualised information about a patient (RCP, 2017). A reliance on NEWS within a similar intervention implemented in northern England was ultimately addressed, with a revised version of the implementation reducing reliance on NEWS while acknowledging the importance of soft signs of deterioration (Russell et al., 2020; Stocker et al., 2021). The Digital Care Home app has used a similar approach, avoiding reliance on NEWS alone and emphasising the importance of soft signs of deterioration and further contextual information. This shows that interventions of this ilk have evolved over time to become better suited to the medical complexity of care home residents and the skills of care home staff.

This qualitative evaluation has raised three issues that could influence future implementations of this and similar interventions. Firstly, despite being part of this implementation’s success, the level of monitoring and staffing required to maintain the ongoing success of the implementation could pose barriers to maintaining current levels of support and the replicability of this intensive implementation process.

Secondly, the belief held by two community nurses that care home staff could be “needy”, and “manipulative” is problematic, suggesting a potential for strained relationships across core stakeholders; a disconnect between care homes’ and community nurses’ understandings concerning expectations and the purpose of the app. Negative views towards care staff may be linked to the stigma and diminished status given to those working in aged care and long-term care facilities (Manchha et al., 2021; Ostaszkiewicz et al., 2016). Implementors of similar interventions should be alert to the potential for such cultural issues to influence implementation success.

Thirdly, digital skills and preparedness in care homes are limited. This issue needs to be addressed, not just for the success of the Digital Care Homes app, but to ensure that care homes are better prepared for future pandemics and digital interventions in general, including interventions designed to support the well-being of residents (Gutman & Shade, 2020; Stadler, 2021; Subramaniam & Woods, 2016; Zamir et al., 2020). The UK Government has pledged to have all care homes digitised by March 2024 (DHSC, 2022). Upholding this pledge appears to be vital to ensuring adult social care can provide a range of person-centred care interventions, and timely and appropriate health care.

### Strengths and limitations

We captured the experiences and views of a range of stakeholders, including care home residents who are often excluded from such evaluations due to challenges of recruitment. Most care home staff interviewed were senior carers, the most common user of the Digital Care Homes application, providing data on the real world, practical experience of the intervention. This is a strength this evaluation.

No care home nurses were recruited and from the NHS perspective we only spoke with community nurses. This may have influenced the findings produced.

### Conclusion

The HealthCall Digital Care Homes app appears to be a feasible, appropriate and legitimate intervention to support improved referral, triage and health care support for non-urgent health care needs of care home residents. The comprehensive implementation process that welcomed feedback to support improvements to the intervention and implementation is the core of this intervention’s success. For this and similar interventions to achieve success nationally, implementations require rapport building and a willingness among those driving the implementation to listen to the views of end users. Ensuring that care homes are digitally enabled with a digitally literate workforce will require structural and economic support from national and local policy makers and care home providers.

## Data Availability

In line with our research approvals and participant consent, data for this work are not publicly available.

## Acknowledgements

The team would like to thank all participants for their time, HDRUK for their funding and support, and the members of the *HDRUK Northern Partnership Care Homes Project Team* involved in this evaluation.

## Conference Presentations

This paper, in draft form, was delivered as an oral conference presentation at the 6^th^ International Long-term care Policy Network (ILPN) Conference in London on September the 10^th^ 2022.

## Conflict of Interest

Authors declare that they have no competing interests.

## Author Contribution

NP, JK, SM were involved from the idea generation phase. SR, RS and ZC drafted study documents, collected data and led the analysis. BH and NP oversaw the qualitative work. SR drafted this paper, with contributions from co-authors.

## Ethics Approval

Ethical approval was granted by HRA and Health and Care Research Wales (ref: 20/LO/0962) and Newcastle University (ref: 12966/2020). The team adhered to accepted ethical standards, ensuring that informed consent was obtained from participants prior to the collection of data. Data were organised in secure shared folders. Only members of the qualitative team were privy to any non-anonymised data. Data were anonymised for analysis and reporting.

## Notes

### Competing Interest Statement

The authors have declared no competing interest.

### Funding Statement

Funded by the Health Data Research UK (HDR UK) Better Care Programme

### Author Declarations

Ethical approval was granted by HRA and Health and Care Research Wales (ref: 20/LO/0962) and Newcastle University (ref: 12966/2020).

## References

Asiedu, G. B., Fang, J. L., Harris, A. M., Colby, C. E., & Carroll, K. (2019). Health Care Professionals’ Perspectives on Teleneonatology Through the Lens of Normalization Process Theory. Health Sci Rep, 2(2), e111. https://doi.org/10.1002/hsr2.111

Barker, R. O., Hanratty, B., Kingston, A., Ramsay, S. E., & Matthews, F. E. (2021). Changes in health and functioning of care home residents over two decades: what can we learn from population-based studies? Age Ageing, 50(3), 921–927. https://doi.org/10.1093/ageing/afaa227

BGS. (2011). Quest for quality: British Geratrics Society joint working party inquiry into the quality of healthcare support for older people in care homes. A call for leadership, partnership and quality improvement. https://www.bgs.org.uk/sites/default/files/content/attachment/2019-08-27/quest_quality_care_homes.pdf

BGS. (2021). Ambitions for change: Improving healthcare in care homes https://www.bgs.org.uk/resources/ambitions-for-change-improving-healthcare-in-care-homes

Brangan, E., Banks, J., Brant, H., Pullyblank, A., Le Roux, H., & Redwood, S. (2018). Using the National Early Warning Score (NEWS) outside acute hospital settings: a qualitative study of staff experiences in the West of England. BMJ Open, 8(10), e022528. https://doi.org/10.1136/bmjopen-2018-022528

Braun, V., & Clarke, V. (2019). Reflecting on reflexive thematic analysis. Qualitative Research in Sport, Exercise and Health, 11(4), 589–597. https://doi.org/10.1080/2159676X.2019.1628806

Braun, V., & Clarke, V. (2021). To saturate or not to saturate? Questioning data saturation as a useful concept for thematic analysis and sample-size rationales. Qualitative Research in Sport, Exercise and Health, 13(2), 201–216. https://doi.org/10.1080/2159676X.2019.1704846

Chu, C. H., Ronquillo, C., Khan, S., Hung, L., & Boscart, V. (2021). Technology Recommendations to Support Person-Centered Care in Long-Term Care Homes during the COVID-19 Pandemic and Beyond. Journal of Aging & Social Policy, 33(4-5), 539–554. https://doi.org/10.1080/08959420.2021.1927620

Dey, I. (1999). Grounding Grounded Theory. Guidelines for Qualitative Inquiry. Academic Press

DHSC. (2022). Policy paper: A plan for digital health and social care. Retrieved from https://www.gov.uk/government/publications/a-plan-for-digital-health-and-social-care/a-plan-for-digital-health-and-social-care

Edelman, L. S., McConnell, E. S., Kennerly, S. M., Alderden, J., Horn, S. D., & Yap, T. L. (2020). Mitigating the Effects of a Pandemic: Facilitating Improved Nursing Home Care Delivery Through Technology [Editorial]. JMIR Aging, 3(1), e20110. https://doi.org/10.2196/20110

Gordon, A. L., Franklin, M., Bradshaw, L., Logan, P., Elliott, R., & Gladman, J. R. (2014). Health status of UK care home residents: a cohort study. Age Ageing, 43(1), 97–103. https://doi.org/10.1093/ageing/aft077

Grant, S. (2018). Limitations of track and trigger systems and the National Early Warning Score. Part 1: areas of contention. Br J Nurs, 27(11), 624–631. https://doi.org/10.12968/bjon.2018.27.11.624

Grant, S., & Crimmons, K. (2018). Limitations of track and trigger systems and the National Early Warning Score. Part 2: sensitivity versus specificity. Br J Nurs, 27(12), 705–710. https://doi.org/10.12968/bjon.2018.27.12.705

Gutman, G., & Shade, M. (2020). Digital Interventions to Promote Health and Well-Being of Older Adults. Innovation in Aging, 4(Supplement_1), 755–755. https://doi.org/10.1093/geroni/igaa057.2721

Harrison, J. K., McKay, I. K., Grant, P., Hannah, J., & Quinn, T. J. (2016). Appropriateness of unscheduled hospital admissions from care homes. Clin Med (Lond), 16(2), 103–108. https://doi.org/10.7861/clinmedicine.16-2-103

Hodgson, P., Greaves, J., Cook, G., Fraser, A., & Bainbridge, L. (2022). A study to introduce National Early Warning Scores (NEWS) in care homes: Influence on decision-making and referral processes. Nurs Open, 9(1), 519–526. https://doi.org/10.1002/nop2.1091

Leonard, M., Graham, S., & Bonacum, D. (2004). The human factor: the critical importance of effective teamwork and communication in providing safe care. Quality and Safety in Health Care, 13(Suppl 1), i85–i90. https://doi.org/10.1136/qshc.2004.010033

Manchha, A. V., Walker, N., Way, K. A., Dawson, D., Tann, K., & Thai, M. (2021). Deeply Discrediting: A Systematic Review Examining the Conceptualizations and Consequences of the Stigma of Working in Aged Care. Gerontologist, 61(4), e129–e146. https://doi.org/10.1093/geront/gnaa166

May, C., & Finch, T. (2009). Implementing, Embedding, and Integrating Practices: An Outline of Normalization Process Theory. Sociology, 43(3), 535–554. https://doi.org/10.1177/0038038509103208

NHSHealthCall. ([online]). HealthCall. Who We Are. Retrieved 06.05.2021 from https://nhshealthcall.co.uk/about/

Ostaszkiewicz, J., O’Connell, B., & Dunning, T. (2016). ‘We just do the dirty work’: dealing with incontinence, courtesy stigma and the low occupational status of carework in long-term aged care facilities. J Clin Nurs, 25(17-18), 2528–2541. https://doi.org/10.1111/jocn.13292

Oung, C., Rolewicz, L., Crellin, N., Kumpunen, S. (2021). Research summary September 2021: Developing the digital skills of the social care workforce, Evidence from the Care City test bed. https://www.nuffieldtrust.org.uk/files/2021-09/workforce-research-summary-final.pdf

RCP. (2017). Royal College of Physicians, National Early Warning Score (NEWS) 2 standardising the assessment of acute-illness severity in the NHS. https://www.rcplondon.ac.uk/projects/outputs/national-early-warning-score-news-2

Ross, J., Stevenson, F. A., Dack, C., Pal, K., May, C. R., Michie, S., Yardley, L., & Murray, E. (2019). Health care professionals’ views towards self-management and self-management education for people with type 2 diabetes. BMJ Open, 9(7), e029961. https://doi.org/10.1136/bmjopen-2019-029961

Russell, S., Stocker, R., Barker, R. O., Liddle, J., Adamson, J., & Hanratty, B. (2020). Implementation of the National Early Warning Score in UK care homes: a qualitative evaluation. British Journal of General Practice, 70(700), e793–e800. https://doi.org/10.3399/bjgp20X713069

Scott, L. J., Redmond, N. M., Garrett, J., Whiting, P., Northstone, K., & Pullyblank, A. (2019). Distributions of the National Early Warning Score (NEWS) across a healthcare system following a large-scale roll-out. Emergency Medicine Journal, 36(5), 287–292. https://doi.org/10.1136/emermed-2018-208140

Smith, P. S.-J. C.; Ariti, C.; Bardsley, M.. (2015). Quality Watch - Focus on: Admissions from Care Homes.

Stadler, T., Burtney, L. (2021). Digital Skills in Adult Social Care: Rapid Evidence Review. https://www.skillsforcare.org.uk/Documents/Misc/Digital-Skills-in-Adult-Social-Care-Rapid-Evidence-Review-Mar21.pdf

Steinskog, T.-L. D., Tranvåg, O., Ciliska, D., Nortvedt, M. W., & Graverholt, B. (2021). Integrated knowledge translation in nursing homes: exploring the experiences of practice development nurses. BMC Health Services Research, 21(1), 1283. https://doi.org/10.1186/s12913-021-07282-7

Stocker, R., Russell, S., Liddle, J., Barker, R. O., Remmer, A., Gray, J., Hanratty, B., & Adamson, J. (2021). Experiences of a National Early Warning Score (NEWS) intervention in care homes during the COVID-19 pandemic: a qualitative interview study. BMJ Open, 11(7), e045469. https://doi.org/10.1136/bmjopen-2020-045469

Subramaniam, P., & Woods, B. (2016). Digital life storybooks for people with dementia living in care homes: an evaluation. Clin Interv Aging, 11, 1263–1276. https://doi.org/10.2147/cia.S111097

Zamir, S., Hennessy, C., Taylor, A., & Jones, R. (2020). Intergroup ‘Skype’ Quiz Sessions in Care Homes to Reduce Loneliness and Social Isolation in Older People. Geriatrics, 5(4), 90. https://www.mdpi.com/2308-3417/5/4/90

